# Knowledge, Attitudes and Acceptance of COVID-19 Vaccine among Pregnant Women in Mbeya Region

**DOI:** 10.1101/2025.02.26.25322935

**Authors:** Revocatus Lawrence Kabanga, Vincent John Chambo, Rebecca Mokeha

**Affiliations:** Department of Obstetrics & Gynecology, Mbeya Zonal Referral Hospital, Mbeya, Tanzania; Department of Obstetrics& Gynecology, University of Dar es salaam-Mbeya College of Health and Allied Sciences, Mbeya, Tanzania; Current address: Department of Internal Medicine, Mbeya Zonal Referral Hospital, Mbeya, Tanzania; Department of Internal Medicine, Mwananyamala Regional Referral Hospital, Dar es salaam, Tanzania

## Abstract

**Background:** The coronavirus was discovered in Wuhan, China, in 2019. WHO declared it a pandemic on March 11, 2020. COVID-19 has caused about 580 million illnesses and 6.4 million fatalities worldwide as of August 8, 2022. Africa reported over 8.7 million cases and 173,063 deaths. East Africa reported 1.39 million cases on July 11, 2022. As of 8 August 2022, 37,865 COVID-19 cases and 841 deaths had been confirmed in Tanzania. To prevent serious illness and death from COVID-19, billions of vaccinations have been delivered. In symptomatic pregnant women, the mortality rate is 70% higher than in non-pregnant women. The effort to control COVID-19 in pregnant women in Tanzania is challenging because only 17.6% of the population is properly vaccinated.

**Objectives:** Assessment of COVID-19 vaccine awareness, knowledge, attitude, and acceptance among pregnant women in the Mbeya region.

**Methods:** A descriptive cross-sectional study was conducted in the Obstetrics and Gynecology department of MZRH. Three scores were calculated for participants’ knowledge, attitude, and acceptance of the COVID-19 vaccination. These scores were compared to many sample factors using binary logistic regression and the chi-square test.

**Results:** The study included 233 pregnant women. Social media provided the most COVID-19 vaccine information to 31.33% of responders. The acceptance of vaccine was low by 38.63%, the knowledge on COVID-19 vaccination was poor in 71.24% and most had negative attitudes on the vaccine (76.82%). Chronic health conditions, high-risk pregnancies, awareness of the COVID-19 vaccine, reliable information sources, preference for natural immunity over vaccination, perceived risks to infants post-delivery, attitudes toward the vaccine, and basic knowledge were strongly associated.

**Conclusion:** Pregnant women exhibited low knowledge, attitude, and acceptance of COVID-19 vaccines. Misinformation about the COVID-19 vaccine causes pause. Education on COVID-19 vaccination is needed to enhance vaccine uptake among pregnant women. This group must comprehend COVID-19 immunization importance, safety, and efficacy.

**KEY TERMS:** **ATTITUDE**−Feeling or way of thinking that affects a person’s behavior/action.

**COVID-19** −Indicates an infection caused by Coronavirus 2019.

**KAA**−Knowledge, Attitude and Acceptance.

**KNOWLEDGE**−Facts, information and skills acquired through experience or education.

**PREGNANCY**−Describe the period in which a fetus develops inside a woman’s womb or uterus.

**VACCINE ACCEPTANCE**−Willingness and readiness to get vaccinated.

**VACCINE HESITANCY**−Delay in acceptance or refusal of vaccine despite the availability of vaccine services.

## Introduction

### Background

Severe acute respiratory syndrome coronavirus 2 (SARS-CoV-2), the RNA virus that causes coronavirus disease 2019(COVID-19), also known as SARS-CoV-2 infection, was first diagnosed in Wuhan, China, in December 2019. Due to the rapidly escalating numbers of new infections outside China in less than three months, COVID-19 was declared a pandemic by the World Health Organization (WHO) on March 11, 2020, in a message delivered by Dr. Tedros Adhanom Ghebreyesus, the WHO Director-General (1).As of August 8, 2022, the WHO recorded almost 580 million confirmed cases of COVID-19, resulting in 6.4 million fatalities across 213 countries.(2).

The COVID-19 pandemic impacted numerous countries globally, including those in Africa. Africa was the last continent affected by the pandemic and was anticipated to be the region where the disease would spread rapidly and exert significant impact. As of August 4, 2022, Africa has recorded over 8.7 million confirmed cases, resulting in 173,063 deaths across 47 countries. (2).As of July 2022, the cumulative number of recorded COVID-19 cases in East Africa was 1.39 million, with Ethiopia and Kenya being the most impacted nations.(2).

The United Republic of Tanzania has also been impacted by the COVID-19 pandemic both economically and socially. The country has experienced three waves of the pandemic, with an increased impact of subsequent waves(3). From 16th March 2020, when the initial case was reported, to 6:24 pm CEST on 8th August 2022, there have been 37,865 confirmed cases of COVID-19 and 841 documented deaths.(2).

The COVID-19 pandemic has caused widespread disease and death globally, with the absence of a vaccination significantly contributing to elevated morbidity and mortality rates. COVID-19 vaccines are now being distributed and made accessible in many different countries.(4).

As of 01 August 2022, over 12.4 billion vaccine doses have been provided globally, with more over 4.88 billion individuals fully immunized. (2). As of August 1, 2022, Africa had received over 924 million vaccination doses and delivered more than 639 million doses.(5)

The United Republic of Tanzania adopted the vaccination strategy, receiving its initial consignment of over one million doses in July 2021. As of July 24, 2022, more than 16.8 million doses had been administered, with over 10.5 million individuals fully vaccinated. (2).

Pregnant women constitute a distinct demographic with an increased susceptibility to COVID-19 morbidity and mortality, possibly experiencing a more severe progression of the disease compared to their non-pregnant counterparts.(6). Symptomatic pregnant women face a 70% elevated risk of mortality compared to their non-pregnant symptomatic peers.(7).Numerous arguments exist regarding the safety and efficiency of COVID-19 immunizations in pregnant women; nonetheless, the COVID-19 vaccination in this group is as successful as in the general population, providing dual advantages to both mothers and newborns.(8).

Despite various initiatives to combat the disease via vaccination, reports indicated that only 17.6% of Tanzanians were fully vaccinated, presenting a significant challenge to the global effort to control the COVID-19 pandemic, as the virus is rapidly mutating in association with successive waves of outbreaks.(2).

Tanzanians, similarly to several other Africans, were perceived as at risk of under-immunization prior to the COVID-19 pandemic, exhibiting low levels of vaccination uptake and trust.(9).Furthermore Pregnant women are known to be at significantly higher risk of severe COVID-19 related complications compared with non-pregnant women. Hence, protecting pregnant women against COVID-19 is critical(7).

Pregnant women may be prone to COVID-19 vaccine disinformation particularly anti-vaccination buzz as well as lack of reliable information due to social marginalization and language barriers; Thus the purpose of this study was to assess knowledge on COVID-19 vaccination, attitudes to COVID-19 vaccines and acceptance towards COVID-19 vaccines among pregnant women in Mbeya region, Tanzania so as to create increased awareness related to the importance of COVID-19 vaccine as essential to expand vaccine utilization(10).

### Problem Statement

The World Health Organization declared COVID-19 as a pandemic on March 11, 2020, impacting over 200 countries.(1).This highly contagious disease has imposed a significant burden on the world, resulting in millions of cases and fatalities.(2). It has also triggered tremendous socio-economic and psychological impacts(11).

Billions of COVID-19 vaccines have already been administered to stop the pandemic(2).However, an effective immunization campaign necessitates sufficient knowledge and a positive attitude toward the vaccine. A lack of knowledge and poor attitude may result in delayed decisions regarding vaccination acceptance or outright refusal, despite the availability and accessibility of immunization services.(12)(13).Disinformation regarding COVID-19 vaccines also appeared to play a major role in vaccination reluctance(14). Raising knowledge on COVID-19 vaccination is one way to boost the uptake(15).

Despite Tanzania ministry of health identification and WHO recommendation, the level of knowledge, attitude and acceptance of COVID-19 vaccines among pregnant women is not well known. A study conducted in Tanzania indicated that COVID-19 vaccine hesitancy was approximately 65%.(14).But this single study is not determining the exact extent of the level of acceptance and didn’t involve pregnant women. Thus this study aimed at assessing the level of knowledge, attitude and acceptance to COVID-19 vaccines among pregnant women in Mbeya region.

## Objectives

### Broad Objective

To assess the awareness on COVID-19 vaccine among pregnant women in Mbeya region.

### Specific Objectives

- To determine the level of knowledge on COVID-19 vaccine among pregnant women in Mbeya region.
- To assess the attitude towards COVID-19 vaccines among pregnant women in Mbeya region.
- To determine COVID-19 vaccine acceptance among pregnant women in Mbeya region.

### Hypothesis

I A lack of knowledge and negative attitudes among pregnant women lead to lower acceptance rate of COVID-19 vaccine in Mbeya region.
II Disinformation regarding COVID-19 vaccine among pregnant women contribute to its lower acceptance rate in Mbeya region.

### Research questions

1. What are the knowledge, attitude and acceptance levels regarding COVID-19 vaccine among these pregnant women in Mbeya region?
2. What are socio-demographic and obstetrics characteristics associated with KAA level of pregnant women towards COVID-19 vaccine in Mbeya region?
3. Which factors could lead these pregnant women towards acceptance regarding COVID-19 vaccination in Mbeya region?

### Significance of the Study

The outcome of this study will determine the KAA levels among pregnant women towards COVID-19 vaccine in Mbeya region as the road to create increased awareness of the importance of vaccination, responding to the hot spots of vaccine hesitancy, promoting vaccine utilization and thence establishing herd immunity as well as alleviating severe pandemic situation and improving maternal and newborn health.

## Materials and methods

### Study Design

An institutional based descriptive cross sectional research study was conducted from 01^st^ September 2022 to 06^th^ September 2022 at MZRH.

### Study area

This study was carried out in Mbeya Zonal Referral Hospital, Obstetrics and Gynecology Section, located in Mabatini Ward, Mbeya Urban District, Mbeya Region, Tanzania. It is located at 08°54ˈ21ˈˈ south and 33°25ˈ59ˈˈ east, surrounded by Itigi ward on the south, Nzovwe ward on the east, Sisimba ward on the north, and Mbalizi road ward on the west. The hospital serves about 8 million people throughout six southern highlands regions: Katavi, Njombe, Rukwa, Ruvuma, Iringa, and Mbeya.

### Study population

The study population comprised of pregnant women who attended antenatal clinic at Mbeya Zonal Referral Hospital.

### Eligibility

#### Inclusion criteria

Pregnant women aged 18 years and above who attended antenatal clinic at MZRH during the study period were included.

#### Exclusion criteria

Pregnant women aged 18 years and above who were seriously ill or unable to hear and read, with mental illness were excluded from the study.

### Sampling procedure and sample size estimation

In this study, the simple random sampling technique was used to select pregnant women.

### The sample size required for this study was determined using a formula of cross sectional study by Kish Leslie(16)

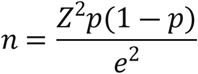

Where

*n* = was the sample size estimated,

P = was the estimated proportion for acceptance of COVID-19 vaccine among pregnant women taken from a study in Ethiopia, p = 18.6%(17).

Z = was the standard normal deviation set at 1.96 which corresponds with 95% confidence interval

e = was the standard error set at 5% marginal error

Then;

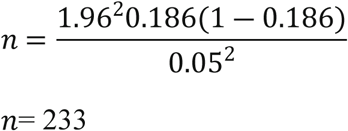

Therefore, the sample size required in this study was 233 participants.

### Data collection tools

Primary data was collected using a structured questionnaire from the target population sampled (pregnant women).

### Questionnaire

The questionnaire consisted of four major sections: sociodemographic data and obstetric features, awareness of COVID-19 vaccination, attitude toward COVID-19 vaccine, and acceptance of COVID-19 vaccine.

### Validity and Reliability

To ensure the study’s validity, a maximum sample size was employed, and the questionnaire was well prepared based on the literature review and reflected study objectives, assessed by the supervisor, and pre-tested before being introduced into the field for data collection.

The reliability of the knowledge, attitude, and acceptance questionnaires was evaluated, with Cronbach’s alpha values of 0.81, 0.82, and 0.92, showing acceptable internal consistency.

The questionnaire was semi-structured and translated into Swahili to prevent misinterpretation of the questions during the interview for optimal reliability.

### Data processing and analysis

The data was validated for completeness, coded, cleaned, and entered into Epi-Data version 7 before being exported to the Statistical Package for Social Science (SPSS) version 27 for analysis. Data were summarized using mean and standard deviation for numerical variables and frequency and percentage for categorical variables. Data were presented in the form of tables, figures, and narratives. The factors related with COVID-19 vaccine uptake among pregnant women in the Mbeya region were investigated using binary logistic regression and the Chi-square test.

### Definition of variables

#### Dependent variables

The basic outcomes of this study (dependent variables) were: - Having good knowledge of COVID-19 vaccine, moderate or poor To assess the participants’ knowledge about the COVID-19 vaccination, nine related questions were posed to them. Each correct answer received a score of one, while incorrect responses received a score of zero. Using Bloom’s cut off point, respondents’ overall knowledge was classed as good if their score was between 80 and 100% (9.0–7.2), moderate if their score was between 60 and 79% (7.1–5.4), and low if their score was less than 60%.(5.4)(18).

Having positive attitude of COVID-19 vaccine, neutral or negative Nine related questions were used to assess the participants’ attitudes toward the COVID-19 vaccination. A correct response received a one-point score, and an incorrect response received zero points. Bloom’s cut off point was used to categorize attitudes towards the COVID-19 vaccine as positive 100-80% (9.0-7.2), neutral 79-60% (7.1-5.4), or negative (less than 60%).(5.4)(19).

Acceptance of COVID-19 vaccine or not Four related questions were asked to the participants, a specific item “If COVID-19 vaccine were recommended for pregnant women, would you get vaccinated?” those who responded “YES” for this question were considered as vaccine acceptance and those who responded “NO” for this question were regarded as vaccine hesitancy(20).

#### Independent variables

These included maternal age, level of education, occupation, marital status, religion, gravidity, parity, gestational age, number of people in the household and number of school age children in the household.

### Ethical Issues

The permission to conduct the research was gained by a formal letter from the UDSM-MCHAS ethical committee, which was presented to the Mbeya area and Mbeya Zonal Referral Hospital administration. With the assistance of the medical professional in charge of RCH, We were able to reach the target population.

Each participant was educated about the research’s purpose, process, benefits, and dangers, and they signed an informed consent form. Participants preserved the right to withdraw from the study at any point during the proceeding of the study. Furthermore, the anonymity of the information revealed was considered by using the information provided solely for research purposes and assigning numbers instead of actual participant names.

### Study Limitations

The study was an institutional cross-sectional study that collected data at a single point in time. Only pregnant women who had access to ANC at the time and agreed to participate were included. Factors of vaccine acceptability may change throughout time.

## Results

### Background characteristics of the participants

The sociodemographic characteristics are summarized in (table 1). A total of 233 pregnant women were interviewed giving a response rate of 100%. The median age of respondents was 30(IQR: 26, 34). One hundred forty-eight (63.52%) were from Mbeya urban district, most of the participants (63.09%) were married, while 110(47.21%) had secondary school education. Compared to other occupations, the majority of participants 57(24.47%) were peasants.

**Table 1.** Showing Sociodemographic characteristics of the sample.

| Variable | Frequency | Percentage (%) |
| --- | --- | --- |
| **District name** |  |  |
| Mbeya urban | 148 | 63.52 |
| Chunya | 43 | 18.46 |
| Mbalali | 22 | 9.44 |
| Kyela | 14 | 6.01 |
| Rungwe | 6 | 2.58 |
| **Maternal age(in years)** |  |  |
| 18–23 | 28 | 12.02 |
| 24–29 | 81 | 34.76 |
| 30–35 | 85 | 36.48 |
| 36–41 | 32 | 13.73 |
| 42–47 | 7 | 3.00 |
| **Marital status** |  |  |
| Married | 147 | 63.09 |
| Others* | 86 | 36.91 |
| **Education level** |  |  |
| No formal education | 22 | 9.44 |
| Primary education | 30 | 12.88 |
| Secondary education | 110 | 47.21 |
| College and above | 71 | 30.47 |
| **Occupation** |  |  |
| HCW | 10 | 4.29 |
| Teacher | 48 | 20.60 |
| Trader | 44 | 18.88 |
| Lawyer | 8 | 3.43 |
| Peasant | 57 | 24.47 |
| Housewife | 25 | 10.72 |
| Unemployed | 41 | 17.60 |
| **Religion** |  |  |
| Christian | 124 | 53.22 |
| Muslim | 105 | 45.06 |
| None | 4 | 1.72 |
| **Number of people in household** |  |  |
| <5 | 139 | 59.66 |
| ≥5 | 94 | 40.34 |
| **Number of children in household** |  |  |
| <5 | 226 | 97.00 |
| ≥5 | 7 | 3.00 |
| *Single, divorced and widowed |  |  |

Health status and obstetric characteristics: One hundred forty (60.09%) of participants were in the third trimester, whereas 203(87.12%) were free of any chronic medical problem. The majority of respondents, 174(74.68%), did not have a high-risk pregnancy. Approximately 67 (28.76%) of the study participants were nulliparous (Table 2).

**Table 2.** Showing obstetric characteristics of the participants.

| **Variable** | **Frequency** | **Percentage (%)** |
| --- | --- | --- |
| **Gestational age(in weeks)** |  |  |
| ≤13 | 7 | 3.01 |
| 14–27 | 86 | 36.91 |
| ≥28 | 140 | 60.09 |
| **Gravidity** |  |  |
| <5 | 212 | 90.99 |
| ≥5 | 21 | 9.01 |
| **Parity** |  |  |
| Nulliparous | 67 | 28.76 |
| <5 | 163 | 69.96 |
| ≥5 | 3 | 1.28 |
| **Chronic medical illness** |  |  |
| No | 203 | 87.12 |
| Yes | 30 | 12.88 |
| **High risk pregnancy** |  |  |
| No | 174 | 74.68 |
| Yes | 59 | 25.32 |

### Source of information on COVID-19 vaccine

The majority of participants (76%) had heard about the COVID-19 vaccine (Fig. 1). Social media was the most trusted source of information on the COVID-19 vaccine, accounting for 31.33% (Fig. 2).

**Fig 1.**
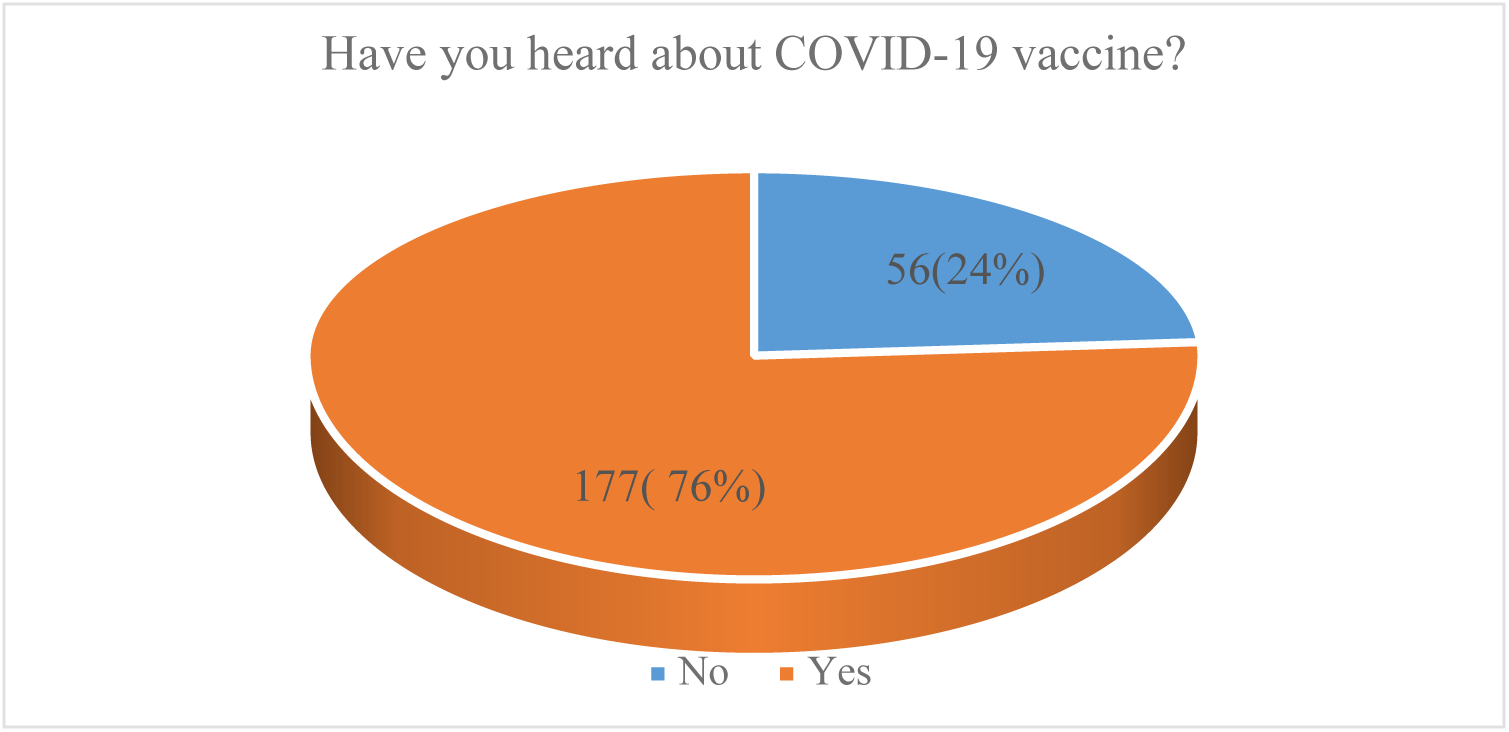
Showing participants who had heard about COVID-19 vaccine.

**Fig 2.**
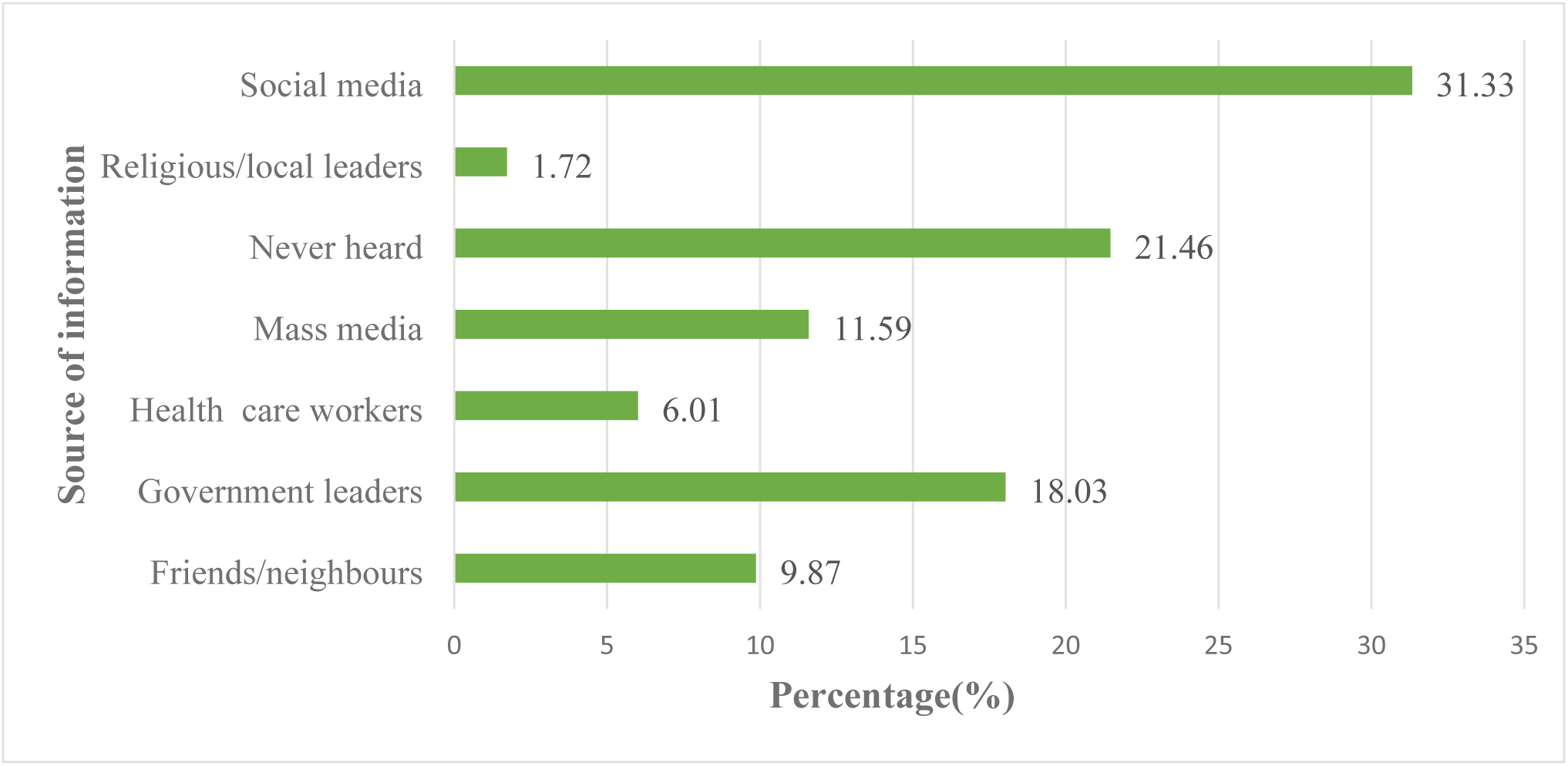
Showing source of COVID-19 vaccine information.

### Knowledge of COVID-19 vaccination

Table 3 presents the knowledge results for the COVID-19 vaccination. The majority of participants (71.24%) had little understanding of the COVID-19 vaccination. When asked if there is a cure for COVID-19, 147 (63.09%) participants responded “Yes,” whereas most participants (75.97%) had heard about the COVID-19 vaccine, and the majority of respondents (76.39%) were aware that COVID-19 immunization had begun in Tanzania.

**Table 3.** Knowledge of COVID-19 vaccine among participants.

| Variable | Frequency | Percentage (%) |
| --- | --- | --- |
| Currently, is there an effective cure of COVID-19? |  |  |
| Yes | 147 | 63.09 |
| No | 86 | 36.91 |
| Have you heard about COVID-19 vaccine? |  |  |
| Yes | 177 | 75.97 |
| No | 56 | 24.03 |
| Is COVID-19 vaccination started in Tanzania? |  |  |
| Yes | 178 | 76.39 |
| No | 55 | 23.61 |
| Are newly discovered COVID-19 vaccine differ from other vaccines? |  |  |
| Yes | 163 | 69.96 |
| No | 70 | 30.04 |
| Do you have enough information about COVID-19 vaccine and their safety? |  |  |
| Yes | 113 | 48.50 |
| No | 120 | 51.50 |
| Can COVID-19 vaccine cause COVID-19? |  |  |
| Yes | 143 | 61.37 |
| No | 90 | 38.63 |
| Do you have enough evidence about COVID-19 vaccine safety during pregnancy? |  |  |
| Yes | 112 | 48.07 |
| No | 121 | 51.93 |
| Do you think COVID-19 vaccine can cause infertility? |  |  |
| Yes | 131 | 56.22 |
| No | 102 | 43.78 |
| Even if there is a vaccine are other preventive measures very important? |  |  |
| Yes | 181 | 77.68 |
| No | 52 | 22.32 |

More than half of participants (51.50%) did not have enough knowledge regarding the safety of the COVID-19 vaccine, while 51.93% did not have enough evidence on the safety of the COVID-19 vaccine for pregnant women. One hundred forty-three (61.37%) respondents indicated that the COVID-19 vaccination may cause COVID-19, while 163 (69.96%) claimed that the newly discovered COVID-19 vaccine differed from existing vaccines. When asked if the COVID-19 vaccine can induce infertility, more than half of the respondents (56.22%) said “yes,” although the majority of participants (77.68%) emphasized the necessity of other COVID-19 prevention methods. Thus, generally, interviewed individuals had “poor” understanding of the COVID-19 vaccination (Fig 3).

**Fig 3.**
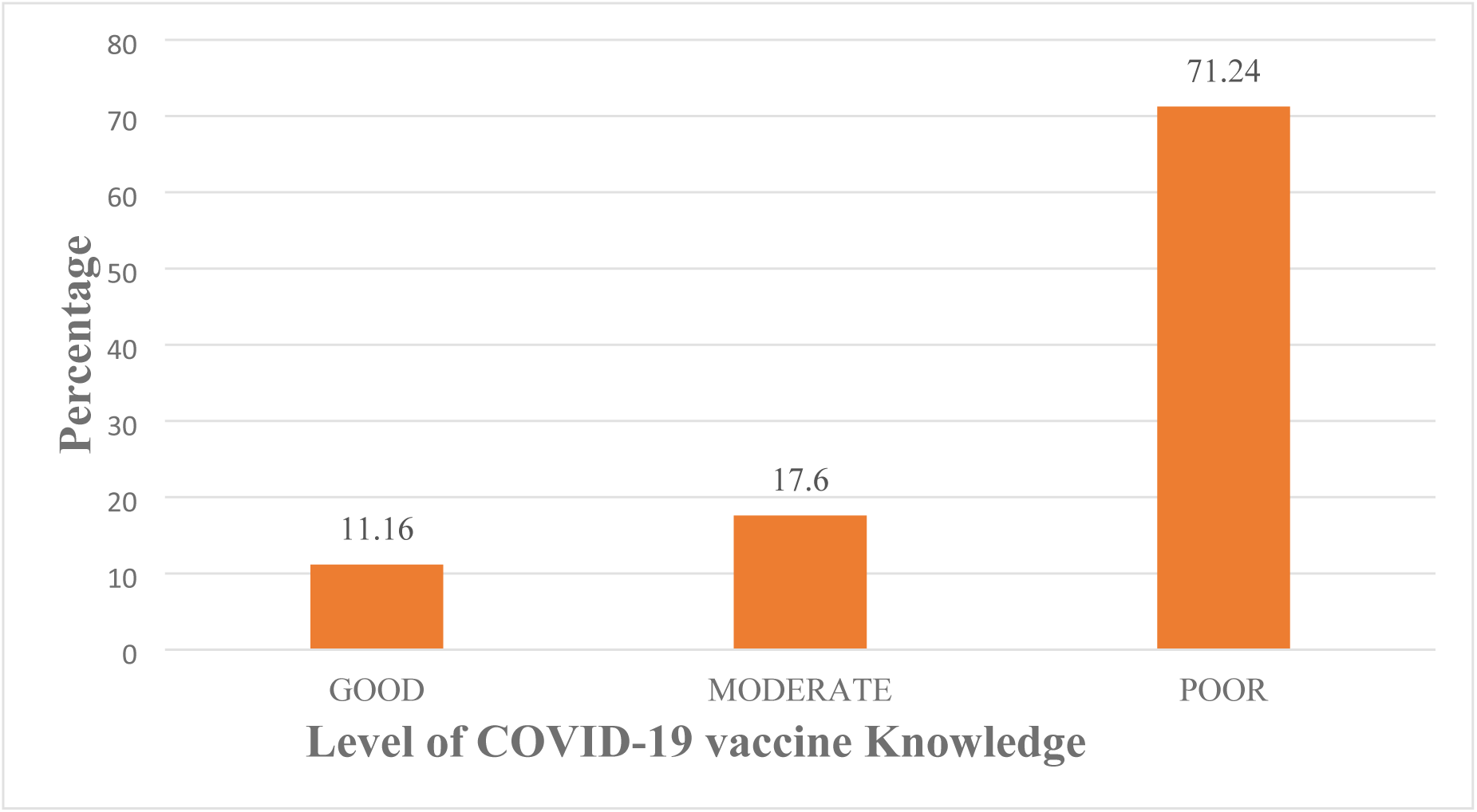
Showing the level of knowledge on COVID-19 vaccine

### Attitude towards COVID-19 vaccine

One hundred and seventy-nine (76.82%) of those interviewed had a negative attitude regarding COVID-19 vaccinations. More over half of the participants (60.94%) believed that the government of the United Republic of Tanzania made decisions in their best interests regarding COVID-19 vaccine. Approximately 37.77% of respondents agreed that the COVID-19 vaccine was necessary for pregnant women, while 66.95% and 58.80% were concerned that the vaccine might harm their bodies and unborn babies, respectively. (table 4)

**Table 4.** Attitudes toward COVID-19 vaccine.

| Variable | Frequency | Percentage (%) |
| --- | --- | --- |
| Do you trust that your government is making decisions in your best interest concerning COVID-19 vaccination? |  |  |
| Yes | 142 | 60.94 |
| No | 91 | 39.06 |
| Do reports you hear/read make you reconsider the choice to take COVID-19 vaccine? |  |  |
| Yes | 138 | 59.23 |
| No | 95 | 40.77 |
| Do you trust pharmaceutical companies to provide credible data on COVID-19 vaccine safety & effectiveness? |  |  |
| Yes | 124 | 53.22 |
| No | 109 | 46.78 |
| Do the benefits of COVID-19 vaccine outweighs reported side effects? |  |  |
| Yes | 105 | 45.06 |
| No | 128 | 54.94 |
| Is COVID-19 vaccine essential for pregnant women? |  |  |
| Yes | 88 | 37.77 |
| No | 145 | 62.23 |
| Do you prefer natural immunity than COVID-19 vaccine? |  |  |
| Yes | 149 | 63.95 |
| No | 84 | 36.05 |
| Do you think COVID-19 vaccine can harm your body? |  |  |
| Yes | 156 | 66.95 |
| No | 77 | 33.05 |
| Do you think COVID-19 vaccine can harm your baby? |  |  |
| Yes | 137 | 58.80 |
| No | 96 | 41.20 |
| Is your decision to get vaccinated driven by your husband/partner? |  |  |
| Yes | 105 | 45.06 |
| No | 128 | 54.94 |

The majority of participants (63.95%) preferred natural immunity over the COVID-19 vaccine; only 27.47% of pregnant women were immunized against COVID-19; however, when asked to reconsider the decision to take the COVID-19 vaccine, more than half of participants (59.23%) agreed to reconsider COVID-19 vaccination. The overall opinion towards the COVID-19 vaccination was judged “negative.”” (Fig 4).

**Fig 4.**
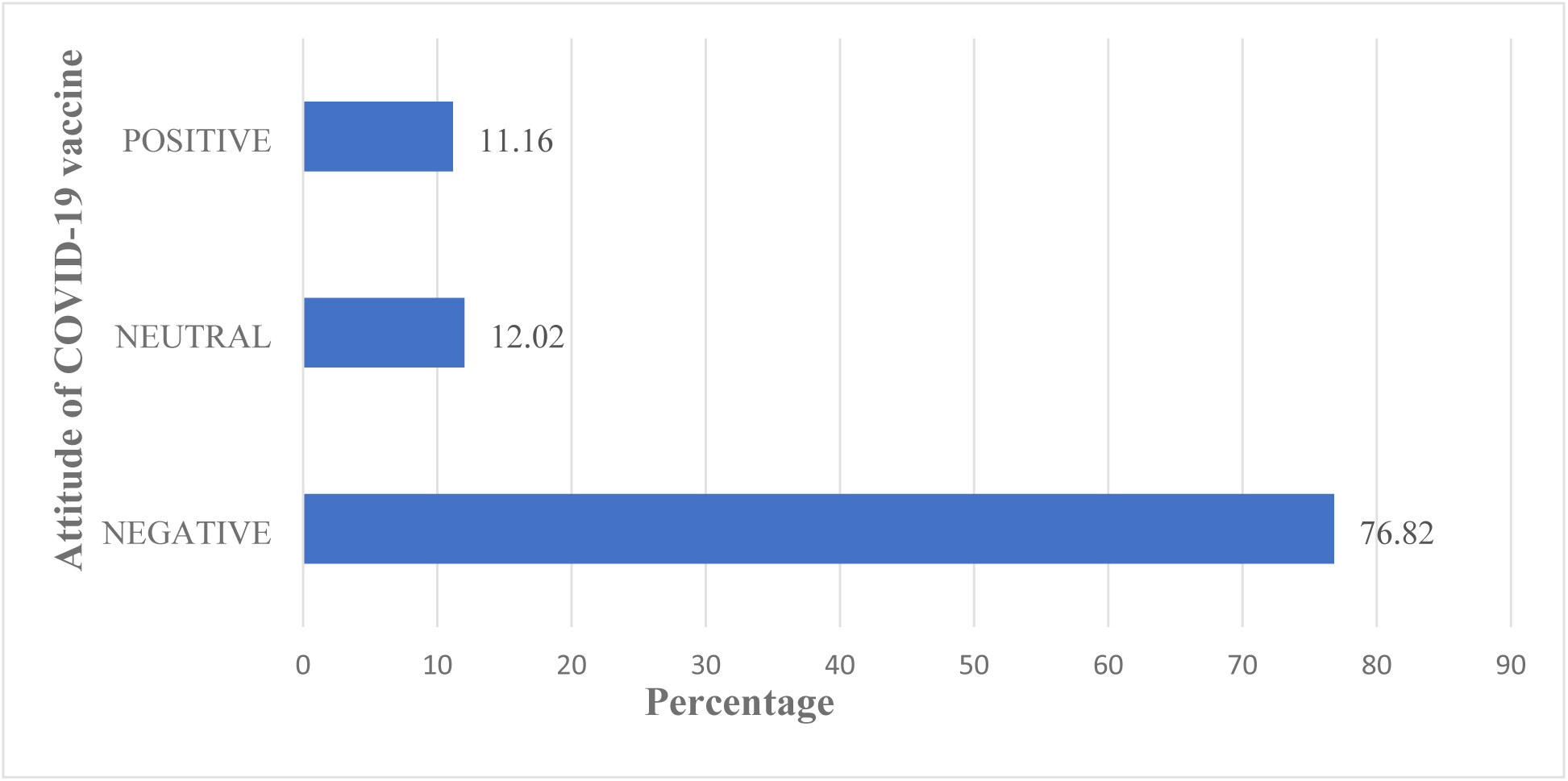
Attitude level towards COVID-19 vaccine.

### Acceptance to COVID-19 vaccine

Ninety-three (39.91%) of the respondents have received additional immunizations within the last five years. Only 27.47% of the pregnant women had already received COVID-19 vaccine, when asked if the COVID-19 vaccine were recommended for pregnant women, will you get vaccinated; only 38.63% of participants were willing to get vaccinated against COVID-19 (Fig 5), while 34.33% of the respondents were willing to have their babies vaccinated after delivery. (table 5).

**Figure 5.**
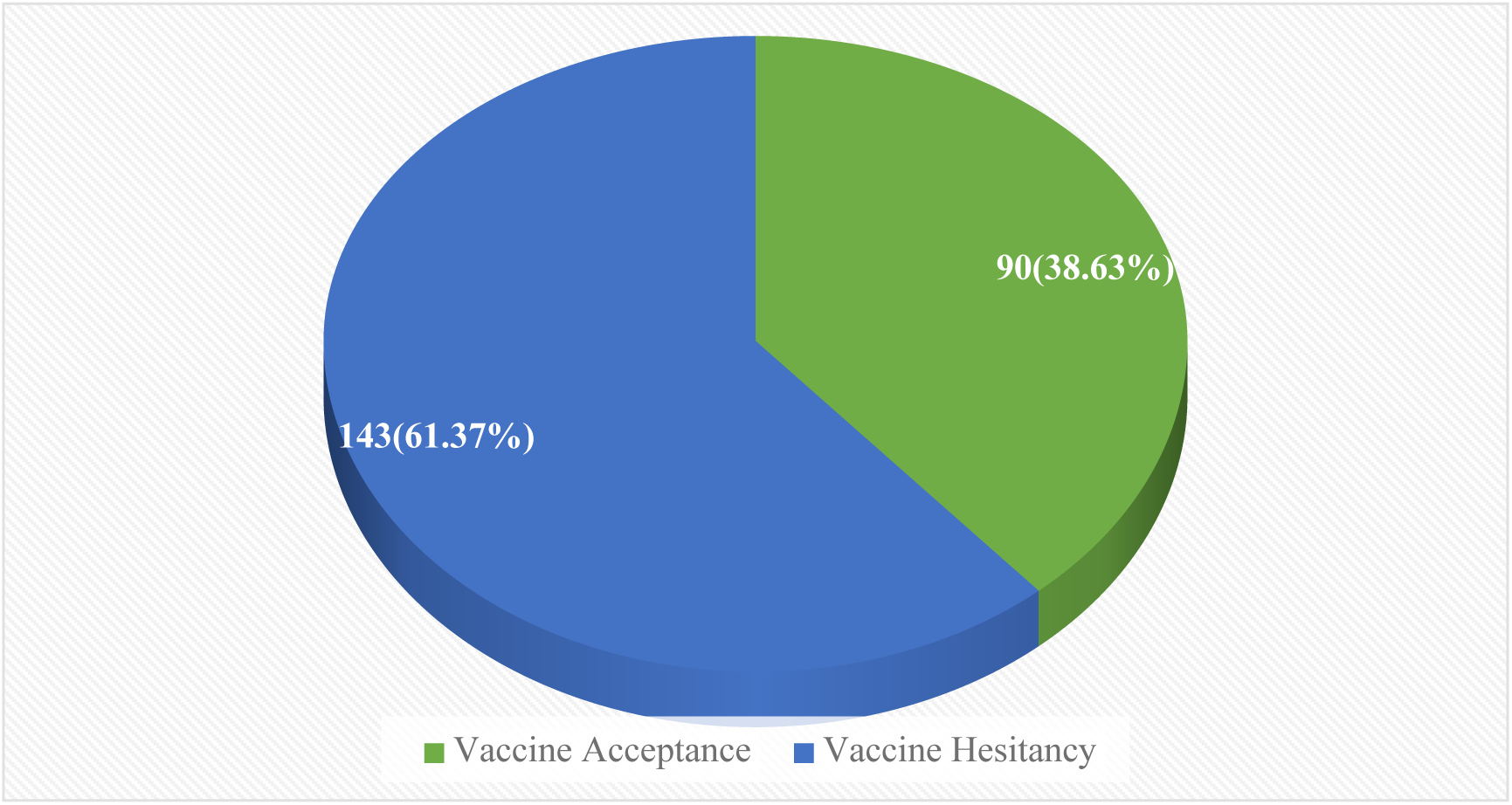
Acceptance level of COVID-19 vaccine.

**Table 5.** Acceptance to COVID-19 vaccine.

| Variable | Frequency | Percentage(%) |
| --- | --- | --- |
| Have you been vaccinated for the last 5 years? |  |  |
| Yes | 93 | 39.91 |
| No | 140 | 60.09 |
| Have you been vaccinated against COVID-19? |  |  |
| Yes | 64 | 27.47 |
| No | 169 | 72.53 |
| If the COVID-19 vaccine are recommended for pregnant women, will you get vaccinated? |  |  |
| Yes | 90 | 38.63 |
| No | 143 | 61.37 |
| Are you going to have your baby vaccinated after birth? |  |  |
| Yes | 80 | 34.33 |
| No | 153 | 65.67 |

### Factors associated with acceptance of COVID-19 vaccine

In binary logistic regression, education level, religion, chronic medical illness, high risk pregnancy, hearing about COVID-19 vaccine, trusted source of information, trusting pharmaceutical companies, preference for natural immunity over COVID-19 vaccine, harm to the baby after delivery, having vaccinated for the past five years, attitude towards COVID-19 vaccine, and knowledge on COVID-19 vaccine were determinants of COVID-19 vaccine acceptability (P-values <0.05).

A chi-square test was also used to investigate parameters linked with the observed KAA for the COVID-19 vaccination. Fisher’s exact test was used to identify significant variables, with a P-value <0.05 indicating statistical significance of the connections.

Poor knowledge of the COVID-19 vaccine was found to be connected with vaccine acceptance, with only 33.13% of pregnant women with low knowledge compared to 48.78% and 57.69% of those with moderate and good knowledge, respectively, having accepted the vaccine. (Chi=7.88, P**=**0.002) (Table 6).

**Table 6.** Relationship between acceptance and COVID-19 vaccine knowledge level.

| Vaccine Acceptance | Level of COVID-19 vaccine knowledge |  |  | Total |  |
| --- | --- | --- | --- | --- | --- |
|  | Good | Moderate | Poor |  |  |
| No | 11 | 21 | 111 | 143 | Key |
|  | 42.31 | 51.22 | 66.87 | 61.37 |  |
| Yes | 15 | 20 | 55 | 90 | Frequency |
|  | 57.69 | 48.78 | 33.13 | 38.63 | Column |
| Total | 26 | 41 | 166 | 233 | Percentage |
|  | 100.00 | 100.00 | 1000.00 | 100.00 |  |

Pearson Chi<sup>2</sup>=7.88 P=0.002

**Table 7.**
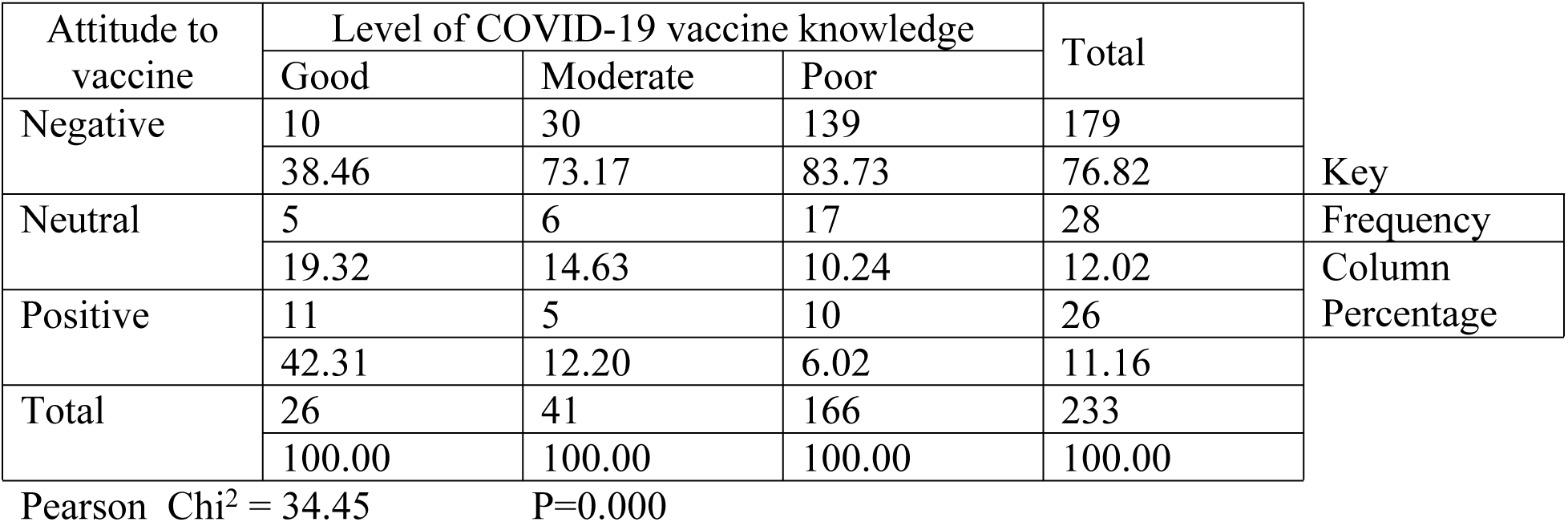
Association between Attitude and COVID-19 vaccine knowledge.

| Attitude to vaccine | Level of COVID-19 vaccine knowledge |  |  | Total |  |
| --- | --- | --- | --- | --- | --- |
|  | Good | Moderate | Poor |  |  |
| Negative | 10 | 30 | 139 | 179 | Key |
|  | 38.46 | 73.17 | 83.73 | 76.82 |  |
| Neutral | 5 | 6 | 17 | 28 | Frequency |
|  | 19.32 | 14.63 | 10.24 | 12.02 | Column |
| Positive | 11 | 5 | 10 | 26 | Percentage |
|  | 42.31 | 12.20 | 6.02 | 11.16 |  |
| Total | 26 | 41 | 166 | 233 |  |
|  | 100.00 | 100.00 | 100.00 | 100.00 |  |

Pearson Chi<sup>2</sup> = 34.45 P=0.000

Furthermore, only 6.02% of participants with poor knowledge of COVID-19 vaccine compared to 12.30% and 42.31% of those displayed moderate and good knowledge respectively had positive attitude towards COVID-19 vaccination (Chi**=**34.45, P=0.000)(Table 8).

**Table 8.**
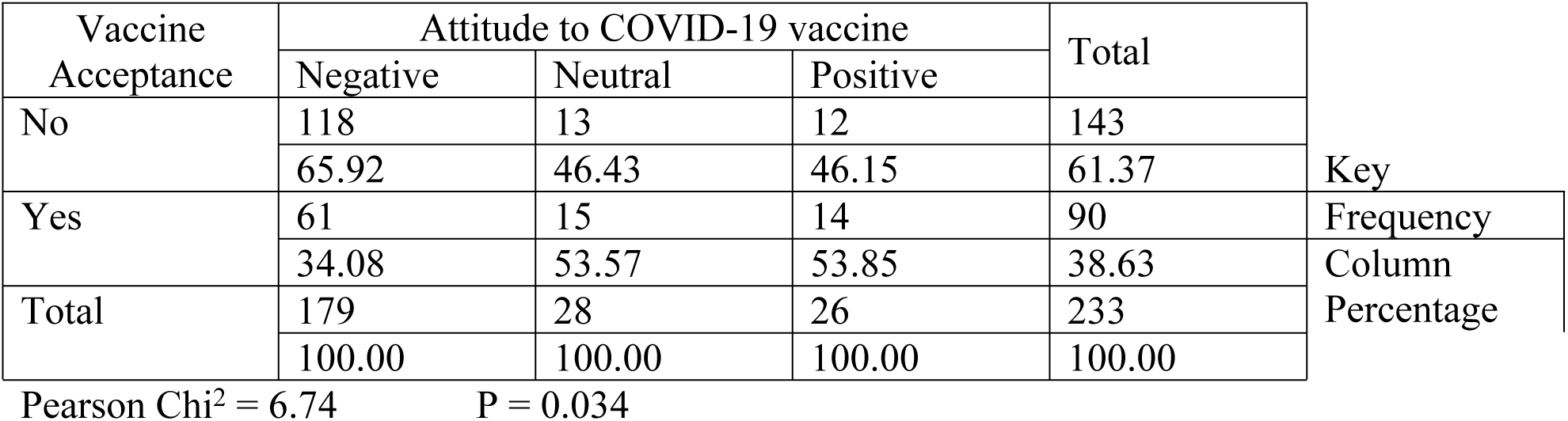
Relationship between Acceptance and Attitude towards COVID-19 vaccine.

| Vaccine Acceptance | Attitude to COVID-19 vaccine |  |  | Total | Key<br>Frequency<br>Column<br>Percentage |
| --- | --- | --- | --- | --- | --- |
|  | Negative | Neutral | Positive |  |  |
| No | 118 | 13 | 12 | 143 |  |
|  | 65.92 | 46.43 | 46.15 | 61.37 |  |
| Yes | 61 | 15 | 14 | 90 |  |
|  | 34.08 | 53.57 | 53.85 | 38.63 |  |
| Total | 179 | 28 | 26 | 233 |  |
|  | 100.00 | 100.00 | 100.00 | 100.00 |  |

Pearson $\chi^2 = 6.74$ P = 0.034

Furthermore, negative attitude toward COVID-19 vaccination was a major determinant in vaccine acceptance, with only 34.08% of pregnant women with negative attitude, compared to 53.57% and 53.85% of those with neutral and positive attitude, respectively, accepting COVID-19 immunization. (Chi=6.74, P=0.034) (Table 8)

## Discussions

This is one of the first studies to report on the level of KAA towards the COVID-19 vaccine and investigate relevant factors influencing COVID-19 vaccine acceptance among pregnant women in Tanzania’s Mbeya Region. The majority of responders had heard about the COVID-19 vaccination. The results show that social media (31.33%) is the most reliable source of information on the COVID-19 vaccination. During the epidemic, users sought the most up-to-date information; however, effective communication is an unavoidable component of COVID-19 crisis response, and attempts to reach the public can take many forms from numerous sources.

This study, like other studies conducted in Tanzania, found that social media platforms are the most efficient means to communicate health-related information about the COVID-19 vaccine due to their extensive functionality.(21). Alternative methods must be employed in remote places with restricted internet connectivity. Reliable and timely communication has been the key to the effectiveness of any control efforts that include the public.(22). Information provided only through social media can be inaccurate, accounting for a decreased vaccine uptake rate among participants (23). Overcoming widespread COVID-19 vaccination hesitation necessitates a concerted public health communication channel on which people can trust, rely, and act. (24) approximately 40.80% of participants had a poor knowledge of the COVID-19 vaccination. Alternatively, a study conducted in Kilimanjaro, Tanzania. (14) demonstrated moderate knowledge of the COVID-19 vaccination. Of note was the response to the question of whether the COVID-19 vaccine can induce infertility, with more than half of respondents (69.10%) saying yes and 30.90% saying no. Misinformation persists, since a significant proportion of responders believe that the COVID-19 immunization can induce infertility.

Furthermore, participants were interviewed about their attitudes toward the COVID-19 vaccine, with 76.82% expressing a negative attitude. This is in line with a study conducted in Tanzania (14), where (65.52%) of participants reported negative attitude towards COVID-19 vaccine. These findings are contrast to a study done in China (25) were 91.3% had positive attitude towards COVID-19 vaccine. The difference may result from study population and profound impact on work, income and daily life as seen among Chinese residents.

A small number of study participants (38.63%) accepted the COVID-19 vaccine. This finding is higher than the study done among pregnant women(18.5%) attending ANC at Debre Markos, Ethiopia(17), but lower than another study done in Ethiopia 62.6%(26) Kuwait(53.1%) (27), Saudi Arabia (90.4%) (28), Middle eastern population (63.2%)(20),Bangladesh (67%) (29) and Iraq (61.7%)(30). The difference may be due to study population and socio-economic status as shown from the study done in Saudi Arabia and Iraq.

Education level, religion, chronic medical illness, high risk pregnancy, having heard about COVID-19 vaccine, trusted source of information, trusting pharmaceutical companies, preference to natural immunity than COVID-19 vaccine, harm to the baby after delivery, having vaccinated for the past five years, attitude towards COVID-19 vaccine and the knowledge of COVID-19 vaccine were the determinants of vaccine acceptance.

## CONCLUSIONS

The COVID-19 pandemic continues to devastate on lives and lively-hoods around the world however COVID-19 vaccine offers a ray of hope for the future. In Mbeya region, Tanzania there is poor knowledge of COVID-19 vaccine and negative attitudes toward COVID-19 vaccine which have resulted in the low vaccine acceptance rates. Disinformation of COVID-19 vaccine appears to be a factor related with vaccine hesitancy.

These findings emphasize that authorities should implement major educational programs, awareness campaigns and disseminate reliable and credible information about COVID-19 vaccine using media, health policymakers, researchers and stakeholders. All concerned bodies should be actively involved to help achieve higher acceptance rates of COVID-19 vaccine among pregnant women. To improve vaccine coverage, it is essential that pregnant women have sufficient knowledge about effectiveness and safety of COVID-19 vaccine.

## Data Availability

No restrictions

## Supporting information

## ABBREVIATIONS

ACOG: American college of obstetrics and gynecology
AJOG: American Journal of obstetrics and gynecology
ANC: Antenatal clinic
CDC: United States Center for Disease Control and Prevention
CEST: Central European Summer Time
COVID-19: Coronavirus disease 2019
EUA: Emergency use authorization
EUL: Emergency use listing
FDA: Food drug administration
FIGO: International federation of obstetrics and gynecology
GMT: Greenwich mean time
HCW: Health care worker
IgA: Immunoglobin A
IgG: Immunoglobin G
IgM: Immunoglobin M
IRCEP: International registry of coronavirus exposure in pregnancy
KAA: Knowledge, attitude and acceptance
mRNA: Messenger ribonucleic acid
MZRH: Mbeya Zonal Referral Hospital
nCOV: Novel coronavirus
PHC: Population and Housing Census
PPE: Personal protective equipment
RCH: Reproductive and child health
RT-qPCR: Real time quantitative polymerase chain reaction SAGE Strategic advisory group of experts on immunization SARS-CoV-2 Severe acute respiratory syndrome coronavirus 2 SPSS Statistical package for Social Science
UDSM-MCHAS: University of Dar Es Salaam Mbeya College of Health and Allied Sciences
UVIKO-19: Ugonjwa wa virusi vya korona 2019
WHO: World Health Organization

## Notes

### Competing Interest Statement

The authors have declared no competing interest.

### Funding Statement

No fund

### Author Declarations

The permission to conduct the research was gained by a formal letter from the UDSM-MCHAS ethical committee, which was presented to the Mbeya area and Mbeya Zonal Referral Hospital administration.

